# Social capital and informational pathways towards sexual and reproductive health and rights literacy among young women in Angola: A cross-sectional study across three provinces

**DOI:** 10.1101/2025.01.14.25320530

**Authors:** Gunilla Elise Priebe, Arciolanda Macama, Francisca Van Dúnem dos Reis, Barbora Kessel, João Ernesto Valles Van Dúnem, José Katito, Nawi Ng, Karin Engström

**Affiliations:** School of Public Health and Community Medicine, University of Gothenburg (UGOT), Box 469, 405 30 Gothenburg, Sweden; Faculty of Economics and Management, The Catholic University of Angola (UCAN), Av. Pedro de Castro Van-Dúnem Loy 24, Luanda, Angola; National School of Public Health, Nova University of Lisbon, Avenida Padre Cruz, 1600-560 Lisbon, Portugal; The Department for Global Public Health, Karolinska Institutet, 171 77 Stockholm, Sweden

**Keywords:** Angola, health equity, sexual and reproductive health information, social capital, young women

## Abstract

**Background:** Social capital is a recognised determinant of health, influencing access to information and formal services in context-dependent ways. In Angola, historical legacies and persistent socio-economic and gendered inequalities intersect with uneven institutional provision, shaping the sexual and reproductive health and rights (SRHR) challenges young women navigate. Where education and access to family planning and maternal health services remain limited, health-related information is central to young women’s ability to protect their health and plan their lives. Although social capital may facilitate engagement with health-related resources, less is known about how its network and trust dimensions relate to access to SRHR information.

**Methods:** Cross-sectional survey data were collected from 2,071 women aged 18–24 years living in Huambo, Luanda, and Lunda Sul. Analyses included province-level descriptive statistics on socio-demographic and SRHR characteristics; exploratory factor analysis of Social Capital Integrated Questionnaire indicators; correlations between social, cultural (literacy), and material (household wealth) capital; and logistic regression models assessing associations with the breadth of SRHR-related information and diversity of information sources.

**Results:** Marked socio-spatial variation was observed in informational, material, cultural, and social resources, with particularly low levels of cognitive social capital (interpersonal and institutional trust). Cultural and material capital were correlated across provinces and were more strongly associated with structural than cognitive social capital. More than two-thirds of participants in Luanda and Huambo reported broader SRHR-related information and more diverse sources, compared with about one-third in Lunda Sul. Information was obtained mainly through family, peers, and health professionals. Formal group involvement was associated with broader information access and more diverse sources, whereas informal network support showed more context-dependent associations. Cognitive social capital showed fewer and less consistent associations.

**Conclusions:** Social capital did not uniformly support access to SRHR-related information. While some forms of social connectedness were associated with broader information access, their importance was shaped by wider socio-structural conditions. Relying primarily on existing social environments or trust-building may therefore be insufficient to narrow informational inequities. Strengthening young women’s pathways to SRHR literacy may require sustained health education within gender-responsive services, alongside structural and relational efforts addressing the conditions associated with low institutional trust.

## Background

### Social capital as a determinant of health and well-being

The relevance of social capital to health and well-being at both individual and population levels is well established in empirical research [1–5]. Higher levels of social capital have been associated with better self-rated health [5–7] and greater life satisfaction [8,9]. Social capital is commonly understood as access to social relationships characterised by trust, reciprocity and cooperation [10–13], through which individuals may gain access to key societal resources, including education and health care [2,14–17].

Beyond material and institutional resources, social capital also shapes access to, interpretation of, and engagement with health-related information [1,18]. The structure and quality of social networks influence not only what knowledge is available, but also how information circulates, is assessed, and becomes meaningful in everyday life [1,4,11–13,18]. In this sense, social capital shapes the relational conditions under which health-related knowledge is encountered and acted upon.

Social capital appears to be particularly salient in contexts marked by socio-economic inequality and limited institutional reach [1,12,14,19], where non-state actors often play an important role in mediating access to services and information [9,15–17,20]. In such settings, reliance on social networks may become especially pronounced, highlighting the importance of understanding how relationships structure access to health-related resources and information [11,14].

### Conceptualising social capital: resources, power, and social position

Conceptually, social capital foregrounds the social embeddedness of individual lives and the ways in which relationships structure opportunities and constraints. Among its diverse intellectual foundations, Pierre Bourdieu’s work has been particularly influential in highlighting how social capital, alongside economic and cultural capital, contributes to individuals’ positions within social space and to the reproduction of power and privilege [21–23].

Cultural capital refers to education, specialised knowledge, and symbolic goods that acquire value through socially recognised practices. Social capital, in contrast, is grounded in durable networks of mutual recognition, through which individuals may access others’ knowledge, skills, and material resources [11,12,21,22]. From this perspective, the value of social capital depends not solely on the existence of social ties, but on the resources circulating within them and on individuals’ relative positions within broader social and institutional structures [11–13,21–23].

Empirical research distinguishes between structural social capital—observable networks and patterns of participation—and cognitive social capital, which reflects perceptions of trust, reciprocity, and shared norms [10,13,24]. Whereas structural social capital concerns enacted social relations, cognitive social capital captures orientations towards these relations. These dimensions are often treated as interrelated, with information exchange, social cohesion, and solidarity commonly understood as key mechanisms through which social capital is realised in practice [10,13,23,25–27].

Social capital is further differentiated according to the social contexts in which relationships are embedded. Bonding ties connect individuals who share familiarity or social position, while bridging ties link actors across social, institutional, or symbolic boundaries [12,13,20]. Relationships situated at different levels of proximity–such as family, community, and formal institutions–are thus associated with distinct configurations of social capital. Linking (or vertical) ties are particularly relevant in contexts of inequality, as they mediate access to institutional authority, formal knowledge, and material resources [9,13,17].

### Social capital, inequality, and context

While there is broad agreement that social capital encompasses both formal and informal networks, the resources they provide, and shared norms of trust and reciprocity, increasing attention has been directed to the ways in which its forms and effects are shaped by historical trajectories, material conditions, and power relations [1,11,12,21,22]. Social capital cannot be presumed to be inherently enabling. Rather, it is embedded within existing social hierarchies and unequal distributions of resources, and may, in certain contexts, contribute to the maintenance of these inequalities [3,28–30].

Empirical studies further demonstrate that social networks do not uniformly function as supportive resources [29–30]. In certain contexts, they may entail obligations, surveillance, or forms of social control, particularly for groups with limited access to alternative forms of capital, including women in low-income settings [31–37]. Relatedly, the use of trust as a universal or sufficient indicator of social capital has been questioned, as expressions of trust are shaped by cultural norms as well as by experiences of inequality, exclusion, and violence [25–28]. These debates raise broader questions regarding the extent to which theories of social capital–largely developed in the global North–can be readily assumed to be universally applicable across diverse historical and social contexts [5,11,12].

Against this backdrop, there is growing recognition of the need to extend empirical research on social capital to underrepresented regions and populations, where systematic evidence remains limited [3,5,14,24]. Examining social capital in such contexts not only addresses empirical gaps but also contributes to more context-sensitive theoretical understandings of how social relationships operate under conditions of structural inequality and uneven institutional provision, thereby informing policies and interventions attentive to existing social structures, relational dynamics, and inequalities in access to health-related information.

### Angola as a context shaping social capital formation and mobilisation

Angola offers a particularly instructive setting for examining social capital beyond its original empirical settings, contributing a new empirical basis for understanding how social environments shape the conditions under which health- and rights-oriented strategies and interventions are developed and implemented. Historically, opportunities for sustained social stability conducive to the accumulation of social capital were constrained by violent and extractive colonial rule, followed by a prolonged civil war [38–44]. Contemporary social relations continue to bear the imprint of these trajectories, with lasting implications for social organisation, institutional reach, and interpersonal trust [39,40,45–50]. Present-day Angola is further characterised by marked inequalities in material resources and gendered entitlements [47–49,51–58], which, in comparable contexts, have been associated with social fragmentation and adverse consequences for health, security, and trust [9,12,28,59,60].

Young people–who constitute more than half of the population [61]–navigate a social landscape shaped by widespread poverty and uneven access to public services and infrastructure [46,47,51–53,62]. While internet-based platforms have expanded opportunities for social connection and information access for some, access remains limited outside the capital, Luanda [63]. In combination with authoritarian forms of political governance, these conditions have been described as constraining civic participation, institutional engagement, and formal avenues for collective action [46,56,57,63–66], thereby shaping both the formation and mobilisation of social capital.

Within this context, religious institutions–most notably Christian churches–have played a significant role in the provision of health and education services, while also functioning as sites of collective belonging and informal social protection [67]. Family and kinship networks likewise remain central sources of emotional, social, and economic support [48,50,68]. These predominantly bonding forms of social capital extend into political and economic life, particularly within the informal sector, where women are disproportionately represented [69–71]. In recent years, civil society initiatives focusing on women’s rights have emerged, particularly in urban areas, some of which draw symbolically on the rural *ondjango* tradition [57], invoking collective deliberation and mutual support grounded in norms of care and reciprocity [57,64,72,73].

### SRHR inequalities and access to SRHR-related information

In addition to civil society initiatives, previous research and official statistics underscore the importance of attending specifically to women’s life situations in Angola [51,56–58,61,74]. Women experience systematic disadvantages across domains central to health equity, including educational attainment, labour market participation, and access to essential health services; gender inequality permeates everyday conditions of well-being as well as broader patterns of institutional responsibility for women’s health needs and rights [51,53,55,58,74,75]. These inequalities are reflected in the high prevalence of child marriage and unintended adolescent pregnancy, persistent limitations in access to family planning, and shortages of medically qualified personnel–particularly providers trained in youth-friendly maternal care–which together are associated with Angola’s high maternal and infant mortality rates by global standards [51,56,75–77].

In contexts characterised by under-resourced state systems and substantial SRHR-related challenges, young women’s access to SRHR-related information constitutes a critical dimension of their life situations, with implications for health, autonomy, and life planning [78–83]. Health literacy research identifies access to comprehensive and trustworthy information as a key precondition for developing the capacities required to make informed decisions, translate knowledge into practice, and navigate health systems effectively [78,84–86]. However, pathways to SRHR literacy are shaped not only by the availability of information but also by the social and structural contexts in which knowledge is produced, communicated, and interpreted [80,84,87,88]. The importance of health literacy is therefore contingent on relational conditions such as social legitimacy, trust, and individuals’ positions within families, communities, and institutional settings [ibid]. These dynamics shape health-related opportunities and condition the forms and potential utility of social contacts in everyday life, with interpersonal relationships functioning as central mechanisms for information exchange and meaning-making [84,85,87,89]. Taken together, these considerations point to the need for empirically examining how different configurations of social capital relate to young women’s access to SRHR-related information in contexts marked by structural inequality.

### Study aim and research questions

Despite this theoretical recognition, empirical evidence remains limited on how different forms of social capital relate to young women’s access to SRHR-related information in contexts such as Angola. To address this gap, this study examines the distribution of different forms of social capital among young women in Angola and their association with access to SRHR-related information. It also considers intersections with cultural and material resources by analysing how dimensions of social capital relate to both the breadth of SRHR-related information accessed and the diversity of information sources from which such information was obtained, reflecting important informational conditions for pathways towards SRHR literacy. By examining variation across SRHR-related topics, information sources, and socio-spatial contexts, the study seeks to clarify how access to SRHR-related information is shaped by the social relations of young women’s everyday lives. Using a study population stratified by province, the study addresses the following research questions:

1. How are socio-economic conditions, social capital, and access to SRHR-related information distributed among young Angolan women?
2. How is social capital associated with cultural and material resources, as indicated by literacy and household wealth?
3. Is social capital associated with the breadth of SRHR-related information and the diversity of information sources?

## Materials and methods

### Study design and study population

This cross-sectional study included women aged 18–24 years residing in selected areas of Angola. It forms part of the broader SADIMA project (*Saúde e Direitos das Mulheres em Angola*–Health and Rights of Women in Angola), a mixed-methods research initiative examining the social determinants of young Angolan women’s sexual and reproductive health and rights (SRHR).

A total of 2,109 women met the inclusion criteria and provided informed consent. Five were excluded due to missing age information, and a further 34 because they did not respond to any social capital items, resulting in a final analytic sample of 2,071 participants. An additional file shows this in more detail [see Additional file 1].

### Setting

Angola gained independence from Portuguese colonial rule in 1975, followed by prolonged armed conflict associated with struggles over natural resources, which ended in 2002 [69]. The country has an estimated population of 35 million, approximately 70% of whom reside in urban areas [52,90]. High levels of natural resource wealth have not translated into broadly distributed living standards. Extreme poverty remains widespread, and national coverage of essential health services remains limited to around 40%. Literacy among young people aged 15-24 is estimated at 80.7% for women and 85.9% for men [52,90].

Three provinces–Luanda, Huambo, and Lunda Sul–were purposively selected to reflect variation in social, economic and demographic conditions. These provinces differ in their colonial and wartime histories, levels of income inequality, fertility patterns, educational attainment, employment opportunities, and health system coverage [51]. Huambo and Lunda Sul are predominantly rural provinces with agricultural potential. Huambo has historically been characterised by smallholder farming and the presence of higher education institutions in the provincial capital, whereas Lunda Sul has experienced more limited investment in infrastructure and social services, contributing to some of the highest urban poverty levels nationally [91,92]. Huambo similarly experiences structural disadvantage, although signs of postwar recovery are evident, including infrastructure development and urban expansion [93]. In both provinces, local cultural institutions retain social significance [94].

Luanda province represents a contrasting context. While rural areas share infrastructural constraints common elsewhere in the country, Luanda City has developed into a highly globalised metropolis and Angola’s principal international hub [41,95]. With over eight million inhabitants, it is the country’s largest urban centre, compared with approximately 450,000 residents in Saurimo (Lunda Sul) and 600,000 in Huambo City. Although Luanda has the lowest poverty prevalence nationally, it contains the largest absolute number of people living in poverty, with pronounced inequalities in peri-urban areas [48,50].

### Recruitment and study size

An initial sample size of 754 was calculated using the most recent Demographic and Health Survey (DHS) data available at the time for women aged 15–19 years [51]. Allowing for an anticipated response rate of 80% and a design effect of 2 yielded a target sample of 1,885 participants. The sample was distributed across provinces in proportion to the cubic root of the population size of selected communes [96].

In the absence of up-to-date household registers, a three-stage sampling strategy was employed to capture socio-economic and urban-rural variation. First, six communes per province were selected using an index derived from principal component analysis (PCA) of five indicators: literacy among 15-24-year-olds, recent use of communication technology, and household access to adequate roofing, sanitation, and safe drinking water [96]. Where access was not feasible, communes in Lunda Sul (n = 2) and Huambo (n = 1) were replaced with others of comparable index values.

Second, within urban communes, data collection sites were identified by randomly selecting blocks from a grid [97], while in rural communes data collection was divided equally between the main village and one randomly selected village. Third, interviewers started from a randomly chosen point within each site and used a “spin-the-bottle” method to determine direction, proceeding household by household until quotas were met [98].

### Data collection

Data were collected between 1 February and 19 May 2022 using a structured questionnaire. In Huambo and Lunda Sul, local CSOs facilitated regulatory clearance and supported the recruitment of data collectors (An additional file shows this in more detail [see Additional file 2]).

Face-to-face survey interviews were conducted by trained female interviewers following a week-long project-specific training covering research ethics, interview techniques, and relevant thematic content. Interviews lasted approximately one hour, with refreshments provided to minimise fatigue. To compensate for lost work time, participants in rural areas received 1 kg of salt and 1 kg of rice. Given limited communication channels and travel distances, this level of compensation was considered unlikely to exert undue influence on participation.

Completed questionnaires were transferred daily to a Microsoft Excel database, anonymised, and stored securely. Data quality checks were conducted daily and after each team’s first week of fieldwork to monitor internal consistency and potential interviewer effects. Although the absence of household registers precluded estimation of the total number of eligible women not approached, participation among those invited was high (95%).

### Data collection instrument development

Survey items analysed in this study were developed between July and December 2021 as part of the broader SADIMA questionnaire, which comprised 150 items organised into five sections: general living conditions, psychological well-being, gender norms, SRHR domains, and social capital. Details on variable construction and source instruments are provided in Additional file 3 [see Additional file 3].

Items were developed through a structured, multi-stage process [55]. Priority themes were identified through a scoping review and key informant interviews with representatives of organisations engaged in women’s health and rights in Angola (n = 25), ensuring contextual relevance while drawing on established scholarship. Validated instruments aligned with the identified domains were then systematically reviewed to inform selection and adaptation [10,99–104]. Items were refined through focus group discussions with young women (n = 20) and piloted with members of the target population (n = 66). Throughout, attention was paid to coherent structure, ethically appropriate sequencing, and clear, respectful language. Idiomatic expressions were added where relevant, and key terms were translated into Umbundu and Chokwe in supplementary reference materials. The final questionnaire was formatted for tablet-based administration using Adobe InDesign, with paper versions available as backup.

### Variables

#### Social capital

The social capital module was developed on the basis of the Social Capital Integrated Questionnaire (SC-IQ) [10], selected for its conceptual robustness, contextual adaptability, and influence on subsequent measurement approaches [100,101]. In line with SC-IQ guidance, items were selected according to theoretical relevance and contextual salience, examined using exploratory factor analysis, and confirmed to represent the intended dimensions of social capital. Details on the factor analysis and other variable constructs are provided in Additional file 3 [see Additional file 3].

Factor analysis identified two dimensions of structural social capital: formal group involvement and informal network support, and two dimensions of cognitive social capital: interpersonal trust and institutional trust.

Formal group involvement captured household-level engagement in formal groups, including type of affiliation (ten options; yes/no for each, e.g. cultural association, neighbourhood or village committee, religious or spiritual group), identification of the most important group, benefits derived from participation (eight options; yes/no, e.g. education or vocational training, health services, transport), intensity of participation, and interaction within and beyond the local community. Minor linguistic adjustments were made during instrument development, and response options relating to group types and perceived benefits were adapted to enhance contextual relevance. Respondents were classified as having formal group involvement if their household was affiliated with a formal group, actively participated in its activities, reported receiving benefits, and the group interacted with other groups [see Additional file 3].

Informal network support measured individual-level access to emotional (number of close friends), economic (number of people able to provide a small amount of money if needed), and emergency support (number of people able to provide urgent assistance), as well as being consulted for assistance (number of people who had sought help from the respondent). Respondents were classified as having informal network support if they reported at least basic engagement across all four domains; engagement in at least three domains including at least one at the highest level of supportive contact; or engagement in at least two domains at the highest level (defined as more than three supportive contacts) [see Additional file 3].

Interpersonal trust was rust was measured using one binary item assessing generalised trust and two Likert-scale items concerning trust in community members and expectations of fair treatment. Respondents were classified as having interpersonal trust if they selected the highest level of trust on any of the three items, or a lower but still positive response option on at least two items [see Additional file 3].

Institutional trust was assessed using a single item adapted following formative feedback indicating potential political sensitivity. Rather than directly asking about trust in government officials, participants were asked to select up to three categories of actors they trusted most from a predefined list, with the option to select fewer categories or indicate an absence of trust. Respondents who selected any state-affiliated actors (health care staff; teachers; police or military personnel) were coded as having institutional trust, reflecting trust in public institutions. The same item also included non-state actors (e.g. family and friends, traditional midwives or healers), which were analysed descriptively but not included as a social capital factor [see Additional file 3].

#### Applied social capital: SRHR-related information access and sources

Within the SC-IQ framework, domains capturing operationalised or applied social capital were reviewed in light of the study objectives and the sociocultural context. Particular attention was given to the information and communication domain [10], given its relevance to young women’s everyday lives. In line with the SC-IQ’s conceptualisation of information and communication as embedded within meaningful life domains, and with health literacy research emphasising both knowledge content and informational environments [78–89], this domain was operationalised through reported access to SRHR-related information along two complementary dimensions: (i) SRHR-related information breadth, and (ii) diversity of SRHR-related information sources. While the latter does not measure communicative engagement directly, a broader range of information sources may indicate broader exposure to SRHR-related communication environments.

SRHR-related information breadth was assessed across five topics: knowledge of the fertile period (correct identification: yes; no); and reported access to information on menstrual health; family planning methods; pregnancy- and childbirth-related risks and care; and domestic violence and available support (all coded yes; no). The wording and topic areas were informed by [102,105]. Information breadth was defined as reporting access to information on three or more of these topics. Diversity of SRHR-related information sources was derived from the latter four topics. For each, respondents indicated the sources from which they had received information (family or friends; traditional midwives or healers; health care staff; community groups, church groups or NGO; school staff; radio, television or internet; or other; all coded yes; no). Source diversity was defined as reporting three or more distinct information sources across the four topics. This was interpreted as indicating exposure to a more heterogeneous SRHR-related information and communication environment. The selected SRHR-related topics and information sources were informed by the 2014 Angola DHS [102] [see Additional file 3].

#### Cultural and material capital

To examine how variations in cultural and material resources correlated with different forms of social capital, indicators of cultural and material capital were included, in line with the analytical orientation of the SC-IQ framework [10]. Literacy (yes/no) was used as an indicator of cultural capital, and household wealth, categorised into tertiles, as an indicator of material capital. Both measures were adapted from the 2014 Angola DHS [102] [see Additional file 3].

#### Other socio-demographic characteristics and living conditions

Additional variables included household food insecurity [103], intimate partner violence (IPV) [104], formal education (high; medium; low), work (permanent; temporary; not working), pregnancy history (never pregnant; first pregnancy ≥18 years; first pregnancy before age 18), residential setting (urban; semi-urban; rural), and family planning use (modern and traditional methods; modern only; traditional only; no methods used), largely adapted from the 2014 Angola DHS [102] [see Additional file 3].

### Statistical methods

Distributions of socio-demographic characteristics, general living conditions, cultural and material capital, SRHR-related information breadth, diversity of SRHR-related information sources, and the dichotomised social capital factors, together with the underlying items for each form of social capital, were summarised as counts and proportions by province [see Additional file 3].

Associations between social, cultural, and material capital were examined using pairwise tetrachoric and polychoric correlations between dichotomised social and cultural capital indicators and trichotomised material capital indicators, stratified by province, with 95% bootstrap confidence intervals (CI) based on 500 iterations. Correlations were considered statistically significant when the CI excluded zero. No adjustment for multiple testing was applied.

Multiple logistic regression models were used to examine associations between social capital factors and the two SRHR-related information outcomes: SRHR-related information breadth and diversity of SRHR-related information sources. Models included all social capital factors, together with indicators of material (household wealth index) and capital (literacy) capital. Results are presented as adjusted odds ratios with 95% confidence intervals (CI). All analyses were conducted using SPSS (version 29.0.0.0) or R (version 4.5.0)[106], including the *psych* package (version 2.5.3), [107,108].

## Results

### Participant socio-demographic characteristics and living conditions

Table 1 summarises participant socio-demographic characteristics and living conditions, with results presented for the total study population and as stratified by province.

**Table 1.** Socio-demographic and reproductive characteristics of the study participants.

| Variable | Category | Luanda<br>n=907<br>% | Huambo<br>n=678<br>% | Lunda Sul<br>n=486<br>% | Total<br>n=2071<br>% |
| --- | --- | --- | --- | --- | --- |
| Formal education | High | 61 | 32 | 19 | 42 |
|  | Middle | 29 | 36 | 32 | 32 |
|  | Low | 11 | 32 | 49 | 26 |
|  | *Missing* | *0* | *<1* | *0* | *<1* |
| Literate | Yes | 86 | 54 | 41 | 65 |
|  | No | 14 | 45 | 58 | 34 |
|  | *Missing* | <1 | 2 | <1 | <1 |
| Work | Yes, permanent | 19 | 18 | 6 | 15 |
|  | Yes, temporary | 32 | 29 | 15 | 27 |
|  | No | 49 | 53 | 79 | 57 |
|  | *Missing* | <1 | <1 | 0 | <1 |
| Pregnancy | Never | 56 | 27 | 21 | 38 |
|  | Yes, ≥18 first pregnancy | 25 | 29 | 26 | 27 |
|  | Yes, <18 first pregnancy | 18 | 44 | 53 | 35 |
|  | *Missing* | <1 | <1 | <1 | <1 |
| Family planning methods | Modern and traditional | 21 | 14 | 7 | 15 |
|  | Modern only | 36 | 26 | 23 | 30 |
|  | Traditional only | 3 | 4 | 3 | 3 |
|  | No method used | 31 | 53 | 64 | 46 |
|  | *Missing* | 10 | 4 | 2 | 6 |
| Intimate partner violence | No | 59 | 55 | 34 | 52 |
|  | Yes | 38 | 44 | 67 | 46 |
|  | *Missing* | 3 | 1 | 0.0 | 2 |
| Household night hunger | No | 72 | 50 | 41 | 57 |
|  | Yes | 28 | 49 | 59 | 42 |
|  | *Missing* | <1 | <1 | <1 | <1 |
| Household wealth tertiles | 3d (better-off) | 61 | 17 | 5 | 33 |
|  | 2d | 33 | 34 | 31 | 33 |
|  | 1st | 5 | 48 | 63 | 33 |
|  | *Missing* | 1 | 2 | <1 | 1 |
| Residential setting | Urban | 94 | 44 | 32 | 38 |
|  | Rural | 6 | 56 | 68 | 37 |

Marked variation was observed across provinces. Luanda was predominantly urban, with 94% of participants residing in urban areas, compared with 44% in Huambo and 32% in Lunda Sul. Participants in Luanda generally reported more favourable socio-economic conditions, while those in Lunda Sul reported more limited circumstances; Huambo consistently occupied an intermediate position.

Literacy levels varied substantially by province, ranging from 86% in Luanda to 54% in Huambo and 41% in Lunda Sul. Across all provinces, reported literacy was lower than the proportion of participants indicating completion of medium or higher levels of formal schooling. Household wealth followed a similar gradient, with 61% of participants in Luanda belonging to the highest wealth tertile, compared with 17% in Huambo and 5% in Lunda Sul.

Pronounced provincial differences were also evident in life-course and relational experiences. The reported prevalence of early pregnancy and IPV was lowest among participants in Luanda (18% and 38%, respectively) and highest in Lunda Sul (53% and 67%), with intermediate levels observed in Huambo (44% and 44%).

### Social capital factors and their underlying items

Table 2 presents the social capital factors identified through exploratory factor analysis, organised into structural and cognitive dimensions, together with their underlying items, for the total study population and by province.

**Table 2.** Social capital factors and underlying items among the study participants.

Overall, a majority of participants reported some level of access to structural social capital, as measured by formal group involvement (63%) and informal network support (67%). Prevalence was similar in Luanda and Huambo but notably lower in Lunda Sul (formal group involvement: Luanda 67%, Huambo 72%, Lunda Sul 43%; informal network support: Luanda 78%, Huambo 68%, Lunda Sul 47%). This pattern was reflected in the underlying items, as household affiliation to formal groups was reported by 86% of participants in Luanda, 85% in Huambo, and 52% in Lunda Sul. Across provinces, religious organisations constituted the predominant form of group affiliation, accounting for approximately 90% of reported memberships. The most frequently cited benefits of group affiliation were access to education (46%) and health services (41%).

A similar provincial gradient was observed for informal network support. Participants in Luanda and Huambo more frequently reported access to emotional or financial support than those in Lunda Sul. In Lunda Sul, close to half of participants reported having no one to turn to for minor financial (44%) or emergency support (41%).

By contrast, reported levels of cognitive social capital followed an inverse pattern. Overall prevalence of interpersonal and institutional trust was low (35% and 28%, respectively), with the lowest proportions reported in Luanda (28% and 15%), followed by Huambo (34% and 46%) and Lunda Sul (49% and 27%). Underlying items capturing institutional trust indicated generally limited trust in state-affiliated actors, including health care staff, school personnel, government officials, and police or military personnel. In Luanda, for example, only 9% of participants reported trust in health care staff, compared with 20% in Lunda Sul. Participants in Huambo reported higher levels of trust in health care staff (34%), teachers (14%), police or military personnel (13%), and CSO representatives (26%).

Beyond the institutional trust factor, family members and friends were the most frequently reported objects of trust across all provinces (89%), with nearly half of participants indicating trust exclusively in this group (results not shown).

### SRHR-related information and information sources

Table 3 summarises reported access to SRHR-related information across thematic areas and the sources from which such information was obtained, stratified by province. In Luanda and Huambo, close to 80% of participants reported broader SRHR-related information and greater diversity of information sources, compared with approximately one third of participants in Lunda Sul.

**Table 3.** Access to SRHR-related information and information sources among the study participants.

Across SRHR-related topics, patterns of reported access were broadly similar, although variation between the provinces was evident. Information on pregnancy- and childbirth-related risks and care was consistently the least frequently reported topic. Approximately two thirds of participants in Luanda and Huambo reported access to such information, compared with around one third in Lunda Sul.

Patterns in reported information sources were broadly similar across provinces. Family members and friends, together with formal institutional actors such as health care staff and school personnel, were the most frequently reported sources, although their relative prevalence varied by setting. Family members and friends were reported as information sources by more than 80% of participants in Luanda and Huambo and by around half in Lunda Sul. Media-based sources were most commonly reported in Luanda (77%), less frequently in Huambo (54%), and infrequently in Lunda Sul (26%). Traditional and civil society actors, including religious organisations, were consistently among the least frequently cited sources across provinces.

### Associations between social, material and cultural capital

Table 4 presents correlations between indicators of cultural and material capital and the different dimensions of social capital. Consistent positive associations were observed between cultural and material capital across all provinces. Associations between these resources and structural social capital were observed primarily for informal network support. In Huambo, no statistically significant association was identified between cultural capital and either dimension of structural social capital.

**Table 4.** Correlations between cultural and material capital and social capital dimensions by province.

With regard to cognitive social capital, interpersonal trust was weakly and negatively associated with cultural capital across all provinces. Institutional trust showed weak but significant negative associations with cultural capital among participants in Huambo only.

### Associations between social capital and SRHR-related information breadth and diversity of information sources

Table 5 presents associations between social capital dimensions and two outcomes: SRHR-related information breadth and diversity of SRHR-related information sources. Together, these outcomes reflect reported access to SRHR-related information across multiple topics and a broader range of information sources. Analyses were based on province-stratified logistic regression models including indicators of structural and cognitive social capital, adjusted for cultural (literacy) and material (household wealth) capital.

**Table 5.**
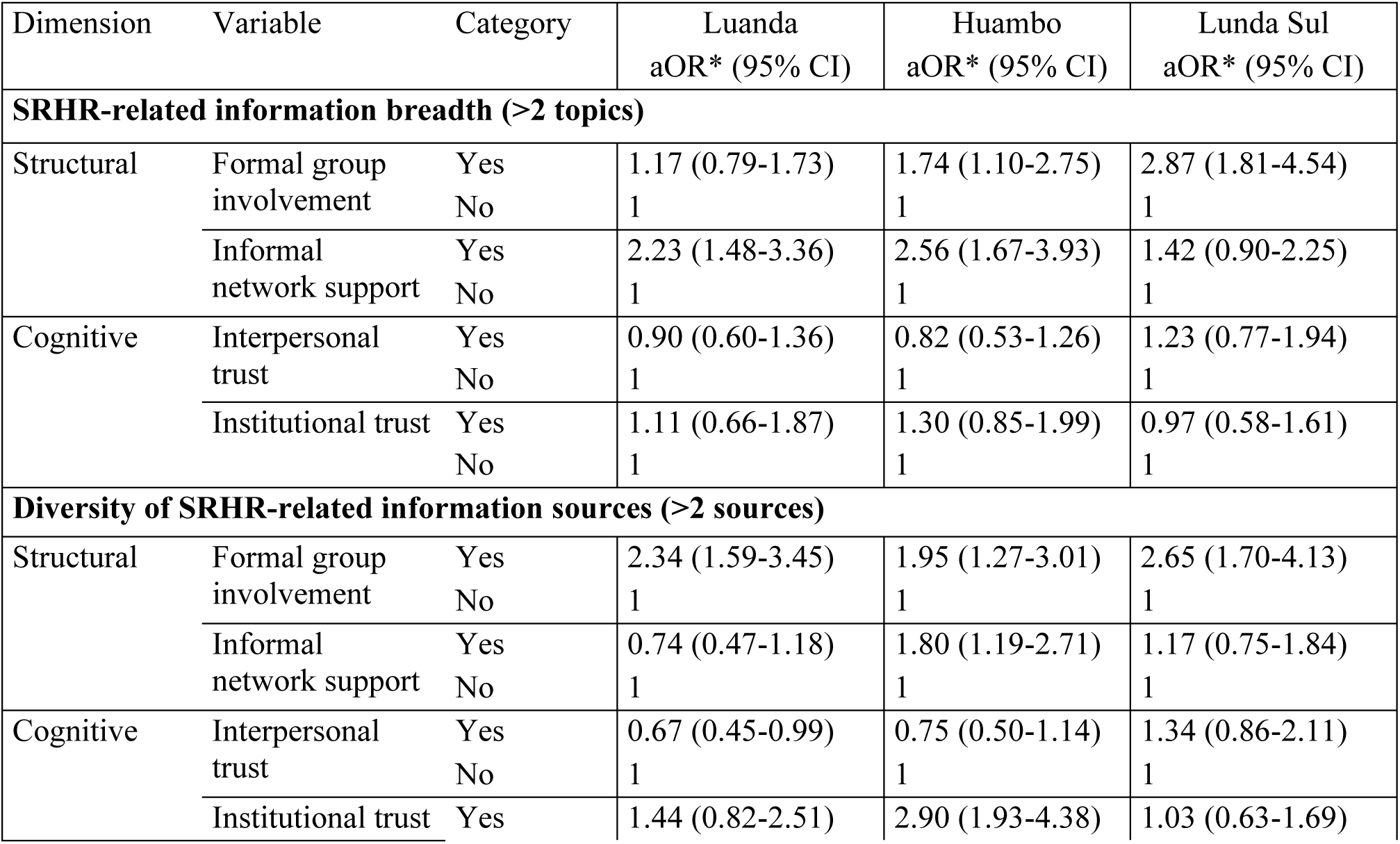
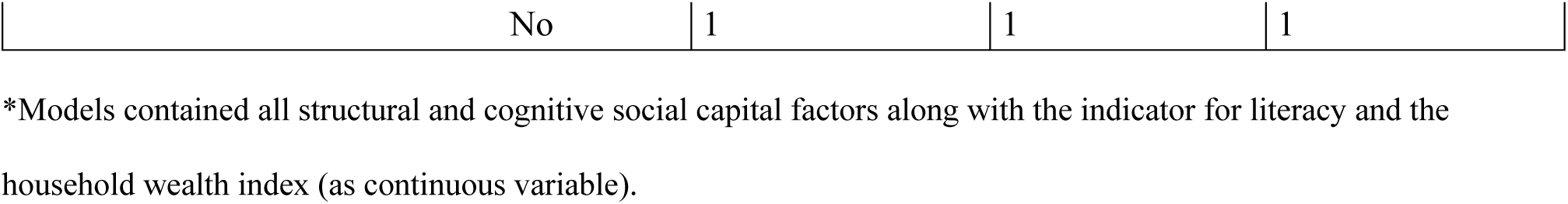
Associations between social capital dimensions and SRHR-related information breadth and diversity of information sources.

Across provinces, structural social capital was consistently associated with SRHR-related information breadth and diversity of information sources, although the magnitude and configuration of these associations varied by province and outcome.

Formal group involvement showed more consistent associations than informal network support. In Huambo, both formal group involvement and informal network support were positively associated with SRHR-related information breadth (formal group involvement: OR 1.74; 95% CI: 1.10–2.75; informal network support: OR 2.56; 95% CI: 1.67–3.93) and diversity of information sources (formal group involvement: OR 1.95; 95% CI: 1.27–3.01; informal network support: OR 1.80; 95% CI: 1.19–2.71).

In Lunda Sul, formal group involvement was associated with SRHR-related information breadth (OR 2.87; 95% CI: 1.81–4.54) and diversity of information sources (OR 2.65; 95% CI: 1.70–4.13). In Luanda, formal group involvement was associated with diversity of information sources (OR 2.34; 95% CI: 1.59–3.45), while informal network support was associated with SRHR-related information breadth (OR 2.23; 95% CI: 1.48–3.36).

Cognitive social capital showed fewer and less consistent associations. No statistically significant associations were observed between interpersonal trust and SRHR-related information breadth. Institutional trust was positively associated with diversity of information sources among participants in Huambo (OR 2.90; 95% CI: 1.93–4.38), whereas a negative association was observed among participants in Luanda (OR 0.67; 95% CI: 0.45–0.99).

## Discussion

### Key findings

This study examined how structural and cognitive dimensions of social capital are associated with young women’s pathways to SRHR literacy, operationalised through SRHR-related information breadth and diversity of SRHR-related information sources across three Angolan provinces with distinct social, economic, and institutional profiles. SRHR literacy was approached through the reported breadth of SRHR-related information and the diversity of information sources, reflecting the informational environments within which young women navigate decisions affecting their health and everyday lives.

Structural social capital and reported access to SRHR-related information were most pronounced in Luanda and Huambo and least evident in Lunda Sul, whereas cognitive social capital followed a different distributional pattern, with the lowest levels observed in Luanda. Across provinces, structural social capital–formal group involvement and informal network support–was positively associated with SRHR-related information breadth and diversity of information sources. Formal group involvement showed the most consistent associations with communication source diversity, suggesting that participation in organised social settings may widen the range of informational context available to young women.

Cognitive social capital–interpersonal trust and institutional trust–displayed fewer and more context-dependent associations. Together, these findings indicate that the development of SRHR literacy among young women is shaped not only by the presence of information, but by the social and institutional environments within which information is shared, interpreted, and made meaningful.

### Social capital as an embedded resource under unequal conditions

The substantial provincial differences observed in literacy, household wealth, employment, and reported exposure to IPV are consistent with previous research and official statistics [51–53]. These inequalities were reflected in the distribution of structural social capital and in SRHR-related information breadth and diversity of information sources. The correlations between cultural and material capital, together with their more modest associations with structural social capital, resonate with Bourdieusian perspectives in which social capital is viewed as intertwined with other forms of capital and shaped by positions within social space [21–23]. For young women, this means that opportunities for support and participation–through which pathways to maternal health literacy develops–are closely tied to their wider living conditions rather than operating independently of them.

**Figure 1.** Proportion of participants with higher levels of social, cultural, and material capital and SRHR-related information breadth and source diversity.

Huambo presented a distinctive configuration. Despite comparatively constrained cultural and material resources, young women in Huambo exhibited structural social capital and SRHR-related information access at levels closer to those observed in Luanda than in Lunda Sul. This highlights how, in specific local contexts, social relations may support young women’s engagement with SRHR-related information, even where formal education and household wealth are more limited.

These findings contribute to ongoing debates about the situated nature of social capital, suggesting that its distribution and potential utility may vary across social fields shaped by historical trajectories and local institutional arrangements [1,12,21–24,32–37]. However, the results also caution against treating social capital as a substitute for material and cultural resources, which remain fundamental to young women’s ability to assess and act upon reproductive health-related information [13,28,109].

### Informal network support and SRHR-related information within close social relations

Associations between structural social capital and material and cultural resources were most evident for informal network support, a form of bonding social capital rooted in close relational environments through which emotional or economic support may be mobilised in times of need [12]. Informal network support was primarily associated with SRHR-related information breadth, rather than with diversity of information sources. While SRHR-related information circulated consistently within family and peer networks, these relational environments did not substantially expand engagement with diverse communication sources, suggesting that informational pathways remained relatively contained within close ties. Where information is concentrated within close ties, opportunities to compare perspectives, question advice, and develop confidence in one’s own judgement may therefore be more limited over time.

Across provinces, family members and friends were the most frequently reported sources of SRHR-related information, whereas engagement with institutional and organisational actors was more uneven and context dependent. The circulation of SRHR-related information was thus largely embedded in relationships characterised by care, reciprocity, and continuity. As previous research has pointed out, these relationships may intersect with locally salient normative expectations concerning sexuality and reproduction, shaping how SRHR-related knowledge is framed, legitimised, and incorporated into everyday SRHR literacy practices [29–37].

These findings underscore that the value of social ties depends not only on their presence but also on the resources embedded within them and individuals’ positions within broader social structures [11,13,17,21,22,37]. Informal network support may therefore facilitate engagement with SRHR-related information while simultaneously reflecting—and in some contexts reinforcing—the unequal conditions under which young women seek, interpret, and use such information. These findings also point to the limits of relying solely on existing social networks to address informational inequalities without attention to the broader conditions under which such networks are formed and accessed.

### Formal group involvement and access to diverse information sources

Formal group involvement showed the most consistent associations with SRHR-related information breadth and diversity of information sources across provinces and was the only social capital dimension associated with both outcomes across strata. Group involvement was strongly linked to religious organisations, reflecting their prominence as sites of collective life and support in Angola [67]. Although churches and community groups were among the least frequently reported direct sources of SRHR-related information, household involvement in such groups was nevertheless associated with informational breadth and communication source diversity.

This pattern suggests that participation in formalised groups may extend young women’s relational reach beyond immediate ties and facilitate indirect pathways to SRHR literacy, even when SRHR-related information does not primarily originate within the group itself. The strongest associations were observed in Lunda Sul, where reported exposure to individual SRHR topics and information sources was lowest. In more resource-constrained settings, formalised group participation may therefore be particularly salient in widening young women’s access to diverse SRHR-related information sources [3,14–16]. Nevertheless, the broader distribution of disadvantage indicates that group involvement alone cannot offset structural and gendered constraints shaping young women’s capacity to engage with and translate available information into practice.

### Trust, institutions, and the differentiated role of cognitive social capital

As an analytically distinct dimension, trust warrants particular attention. Its suitability as a measure of social capital has long been debated [12,13,19,25,27,28]. Given the role trust may play in how reproductive health and rights are negotiated within informal relationships and formal institutions–particularly in contexts marked by pronounced gender and economic inequalities–it remains analytically relevant as a context-dependent feature [6,11,12,26,31–33,37]. At the same time, the findings underscore the limitations of trust as a stand-alone indicator, as it did not consistently emerge as closely linked to structural social capital [6,11–13,25,26]. The absence of positive associations between interpersonal trust and structural social capital aligns with earlier research showing that network participation may develop independently of trust [6,13,25,26].

This divergence was particularly evident in Luanda. Despite being the most materially advantaged setting, 13% of participants reported that they trusted no one, and levels of trust in community actors and formal institutions were the lowest observed in the study, while trust in family members and friends remained comparatively high. This pattern is consistent with earlier characterisations of Luanda city as a socially heterogeneous urban environment marked by uneven institutional accessibility, political tensions, and substantial in-migration from more resource-constrained regions [48,49,110]. The social fragmentation and everyday uncertainty associated with these conditions may be reflected, in part, in the low levels of generalised and institutional trust reported by young women navigating these environments. At the same time, young women in Luanda reported comparatively broad access to SRHR-related information, including through media and internet-based sources, CSOs, and other formalised groups. This suggests that pathways to SRHR literacy may be supported through multiple relational and institutional spaces, even where trust beyond close ties is limited and interactions outside family and friendship networks are approached with caution [6,11–13,15,16,37].

A contrasting configuration emerged among young women in Lunda Sul. Despite more adverse living conditions and more limited SRHR-related information breadth and diversity of information sources, the presence of interpersonal trust was comparatively higher. Rather than signalling expanded practical opportunities, such trust may reflect reliance on close relational ties in contexts where alternative sources of support are scarce and livelihoods are constrained [11,12,31–33,37]. These trust dimensions did not, however, translate into expanded informational pathways, highlighting the limits of interpersonal trust in the absence of supportive institutional and informational infrastructures. This interpretation is reinforced by the very low levels of institutional trust reported in Lunda Sul, which may reflect historically grounded scepticism towards formal institutions in contexts characterised by intermittent state presence and limited institutional reliability and responsiveness [91,92,110]. In settings shaped by extractive economies, such as diamond mining, and without commensurate investment in public services, trust in formal institutions may be less likely to develop as a meaningful social resource [12,13,21,24,38,46,47].

Findings from Huambo point to a different configuration, where a degree of institutional trust coexisted with widespread poverty and predominantly rural conditions. Among young women in Huambo, relatively higher levels of trust in institutional representatives–including health care workers, teachers, and security personnel–were observed. This pattern is consistent with social capital theory emphasising the role of local governance and institutional embeddedness in shaping institutional trust as a relational resource [2,11,12,15,16], and suggests that trust in formal institutions may support engagement with SRHR-related information and services that might otherwise be difficult to approach. For young women with limited material and social resources, trust–understood as an expectation of reciprocity and support–may signal the relational conditions under which seeking advice, assistance, or services becomes feasible.

Taken together, patterns of interpersonal and institutional trust appear to reflect broader conditions of inequality, institutional accessibility, and social change, operating partly independently of participatory or network-based dimensions of social capital [11,12,19,28,38]. These configurations nonetheless shape how young women negotiate relationships, engage with institutions, and assess the credibility and relevance of SRHR-related information in everyday decision-making [6,13,25,26].

### Limitations and generalisability

This study highlights several dimensions of social capital that warrant further theoretical and empirical elaboration, particularly regarding how trust is experienced across diverse living contexts.

A modified version of the SC-IQ [10] was employed to enhance contextual relevance while maintaining theoretical coherence. While this allowed a broader mapping of social capital indicators than is typical in comparable studies [1,5–8,19,24,28,109], it also exposed conceptual and measurement challenges. In line with many previous investigations, neighbourhood relations were grouped with close social ties and treated as bonding social capital [6,10,13,25,26]. The uneven distribution of trust observed—particularly the lower levels reported beyond family and friendship networks—suggests that this categorisation warrants further scrutiny in contexts such as Angola. More fine-grained operationalisations are needed to clarify which social spheres young women perceive as trustworthy and reciprocal, and how these perceptions shape the meaning and function of trust. This is particularly important given that bonding social capital is the form most consistently associated with health outcomes [1,6,14,59].

Linking social capital, operationalised through institutional trust, also proved methodologically challenging. The theoretical understanding of this dimension was largely developed in settings where institutional relationships are assumed to be clearly delineated and openly expressible, which may limit its sensitivity to dynamics in politically constrained environment [24,10–12,28]. In Angola, informal networks are often intertwined with political affiliations and access to public services [62,64,66,69,70], complicating distinctions between bonding, bridging, and linking forms of social capital. Further conceptual and methodological refinement is therefore needed in similar contexts.

The study included only young women and cannot determine whether observed patterns of trust and social capital are specific to this group. The focus on a single age group and the use of a non-random sample limit statistical generalisability. However, the sociodemographic profile of the sample aligns with national data [51], supporting the broader relevance of the findings. While configurations of social capital may vary across provinces, participants share structural conditions characteristic of settings marked by enduring socioeconomic and gender inequalities. The findings may therefore offer insights relevant to comparable contexts.

The outcome measures captured self-reported access to SRHR-related information rather than its quality, depth, or practical applicability. Uneven reporting across subject areas—particularly in relation to pregnancy and childbirth-related complications—indicates the need for future research to examine not only informational reach but also content and usability [80,83,84,88]. Interview-based data collection enabled inclusion across literacy levels and geographical areas but may have introduced social desirability bias. Household wealth indicators should be interpreted cautiously due to skewed provincial distributions, although overall patterns were consistent with national data [51] (see Additional file 3).

Finally, qualitative research would be valuable in deepening understanding of how young women experience, negotiate, and mobilise different forms of social capital in contexts marked by socio-economic and gender inequalities. Such approaches could inform the development of measurement tools with greater cultural and contextual sensitivity.

## Conclusion

This study demonstrates that young women’s access to SRHR-related information in Angola–understood as an important pathway towards effective SRHR literacy–is shaped by context-specific configurations of social capital embedded within broader social, material, and institutional conditions. Trust, in particular, did not function as a stand-alone resource for improving access to SRHR-related information. In this light, the generally low levels of trust observed serve as a reminder that expectations of improved SRHR-related outcomes or informed decision-making among young women may be difficult to sustain in the absence of social and institutional environments characterised by trust, within which sensitive issues can be discussed and reflected upon without fear of sanction or stigma.

While cognitive dimensions of social capital showed few and context-dependent associations, structural social capital–especially formal group involvement–was consistently associated with broader access to SRHR-related information and a greater diversity of information sources. Formal group involvement, particularly in resource-constrained and rural settings, appeared to extend informational pathways beyond immediate social circles, thereby widening the range of contacts through which young women may encounter and discuss SRHR-related information.

Family and peer networks emerged as central to young women’s informational lives, frequently serving as the primary sources of SRHR-related information. Informal network support facilitated access to a broad range of SRHR-related topics; however, it was not associated with exposure to a wider diversity of information sources. This pattern highlights the dual role of bonding social capital as both supportive and potentially constraining in shaping informational pathways to SRHR literacy. More broadly, the findings illustrate that social capital operates cumulatively rather than compensatorily, tending to reinforce existing advantages linked to literacy, household wealth, and access to institutions, rather than offsetting structural inequalities that shape young women’s opportunities to seek, interpret, and act upon health-related knowledge.

From a policy and practice perspective, these findings point to the importance of strengthening equitable, accessible, and youth-responsive SRHR services as a foundation for improving information access. Approaches that rely primarily on existing social networks or trust-building initiatives, without parallel investment in institutional capacity, risk reproducing informational inequalities. In institutionally dense settings, linking young women to schools, health services, CSOs, and reliable media platforms may help broaden informational pathways beyond family and peer networks. In more resource-constrained or rural contexts, targeted engagement with community and faith-based groups may support the dissemination of trustworthy health information, although such efforts cannot be substituted for sustained investment in public services and communication infrastructure.

By situating social capital within young women’s lived realities and institutional environments, this study contributes to a more context-sensitive understanding of how pathways to SRHR literacy are shaped under conditions of inequality, highlighting both the potential and the limits of relational resources for advancing health equity.

## Data Availability

Data is made available upon reasonable request. The data set is not presently available on an external platform, but are included in the journal submission system and will be available when accepted for publication.

## List of abbreviations

CI: Confidence Interval
CSO: Civil Society Organisations
DHS: Demographic Health Survey
IPV: Intimate Partner Violence
PCA: Principal Component Analysis
SADIMA: **SA**úde e **DI**reitos das **M**ulheres em **A**ngola (Health and Rights among Women in Angola)
SC-IQ: Social Capital Integrated Questionnaire
SRHR: Sexual and Reproductive Health and Rights

## Declarations

## Ethics approval and consent

The study adhered to the ethical principles of the Declaration of Helsinki [113]. Approval was obtained from the Ethics Review Board of the Angolan Ministry of Health (24/C.E./2021), the Ethics Review Board of the Universidade Católica de Angola (Approvação 153, CEIH 230), and the Swedish Ethics Review Authority (Dnr 2022-06393-01). Verbal informed consent was secured from all participants in line with DHS procedures [111], appropriate in contexts with high illiteracy. Participants could decline to answer any question. The principles of beneficence, respect for autonomy, fairness, and non-maleficence guided the entire research process.

## Consent for publication

Not applicable

## Availability of data and materials

All data generated or analysed during this study are included in this published article and its supplementary information files.

## Authors’ contributions

GP, as principal investigator, coordinated and implemented the SADIMA project. GP, AM, JEVD and NN designed the study. The questionnaire was developed by GP, AM, FVDR, JK and NN, who also prepared ethical applications. GP, AM, FVDR and NN designed and supervised data collection. GP, BK, JEVD and KE contributed to data management. BK conducted statistical analyses with input from GP and KE. All authors contributed to the interpretation of findings, discussion and conclusions. GP, BK and KE drafted the manuscript with input from all co-authors, who approved the final version.

## Acknowledgements

We would like to express our warmest gratitude to Cidália Gomes and Juliana Tchimoko (ADRA Huambo), Maria Malomalo and Juliana António (Mwana Pwo Lunda Sul), and Francisco Quemba and Yara Sicato (UCAN Luanda) for assuming operational responsibilities during the data collection, Francisco Quemba (UCAN) for digitising the questionnaire, Professor Lucie Laflamme (Karolinska Institute) for her input on the methodology and the draft manuscript.

